# Impact of Reduced Sampling Rate on Accelerometer-based Physical Activity Monitoring and Machine Learning Activity Classification

**DOI:** 10.1101/2020.10.22.20217927

**Authors:** Scott R Small, Sara Khalid, Paula Dhiman, Shing Chan, Dan Jackson, Aiden R Doherty, Andrew J Price

## Abstract

**Purpose:** Lowering the sampling rate of accelerometer devices can dramatically increase study monitoring periods through longer battery life, however the validity of its output is poorly documented. We therefore aimed to assess the effect of reduced sampling rate on measuring physical activity both overall and by specific behaviour types.

**Methods:** Healthy adults wore two Axivity AX3 accelerometers on the dominant wrist and two on the hip for 24 hours. At each location one accelerometer recorded at 25 Hz and the other at 100 Hz. Overall acceleration magnitude, time in moderate-to-vigorous activity, and behavioural activities were calculated using standard methods. Correlation between acceleration magnitude and activity classifications at both sampling rates was calculated and linear regression was performed.

**Results:** 54 participants wore both hip and wrist monitors, with 45 of the participants contributing >20 hours of wear time at the hip and 51 contributing >20 hours of wear time at the wrist. Strong correlation was observed between 25 Hz and 100 Hz sampling rates in overall activity measurement (r = 0.962 to 0.991), yet consistently lower overall acceleration was observed in data collected at 25 Hz (12.3% to 12.8%). Excellent agreement between sampling rates was observed in all machine learning classified activities (r = 0.850 to 0.952). Wrist-worn vector magnitude measured at 25 Hz (Acc_25_) can be compared to 100 Hz (Acc_100_) data using the transformation, Acc_100_ = 1.038*Acc_25_ + 3.310.

**Conclusions:** 25 Hz and 100 Hz accelerometer data are highly correlated with predictable differences which can be accounted for in inter-study comparisons. Sampling rate should be consistently reported in physical activity studies, carefully considered in study design, and tailored to the outcome of interest.

## Introduction

Accelerometers are increasingly used to obtain exposure measures in large observational physical activity healthcare studies [1–3] and to obtain outcome measures in randomised controlled trials [4]. Seven days of physical activity measurement is a common measurement window across a variety of accelerometer studies and provides a suitable representation of individual activity in most scenarios [5–8]. However, in the case of activity monitoring in clinical populations recovering from trauma or a surgical intervention, extended monitoring well beyond a standard seven day protocol may be desired [9]. Extended longitudinal protocols incorporating measurement windows beyond a few weeks requires frequent participant interaction for battery charging, which is burdensome and undesirable [10]. In these cases, a reduction of sampling rate would offer the potential for longer continuous physical activity measurement, while reducing patient and caregiver interactions with the monitoring device.

Currently, Brønd and Ardivsson offer the only side-by-side comparison of accelerometers recording at different sampling rates, recording laboratory-based activities with hip-mounted Actigraph GTX3+ devices [11]. Other studies generate downsampled data from a single accelerometer to assess the effects of varying sampling rates, while relying on the untested assumption that this postprocessing accurately reproduces the recording behaviour of the underlying hardware [12,13]. In addition, studies of this nature have been limited to the ActiGraph GTX3+ and GENEA triaxial accelerometers [11–14], and not the Axivity AX3 which has been used in the UK Biobank study of 100,000 participants [1]. This dataset is a valuable resource and can serve as a reference for physical activity within a wide range of clinical populations. Therefore, the ability to compare physical activity data between studies integrating non-standard sampling rates remains unverified.

The aim of this study was to validate data collected at a reduced sampling rate (25 Hz) in comparison to that collected at the commonly used rate of 100 Hz within the AX3 accelerometer. The objectives were to: 1) identify any effect of sampling rate on vector magnitude both overall and for specific free-living activities; 2) characterise the effect of reduced sampling rate on machine learning activity classification; 3) develop a transformation so that data collected at standard and reduced sampling rates can be directly compared.

## Methods

### Study design

Ethical approval for participant recruitment was obtained from the Central University Research Ethics Committee of the University of Oxford (Ref: R63137/RE001). Written informed consent was obtained from adult volunteers (aged 18 and above) with no lower limb injury within the previous 6 months and who were able to walk without an assistive device. Participants were recruited through advertising within Oxford University, the local community and in senior citizen groups.

Participants were instructed to wear four triaxial accelerometers (AX3, Axivity, Newcastle, UK) for 24 hours, except during bathing or swimming activities. Participants could remove sensors at night if they disrupted sleep. Two accelerometers were placed side-by-side on the dominant wrist using a wristband, and two on the dominant-side hip, waist level at the anterior-posterior midline via a belt clip (Figure 1).

**Figure 1:**
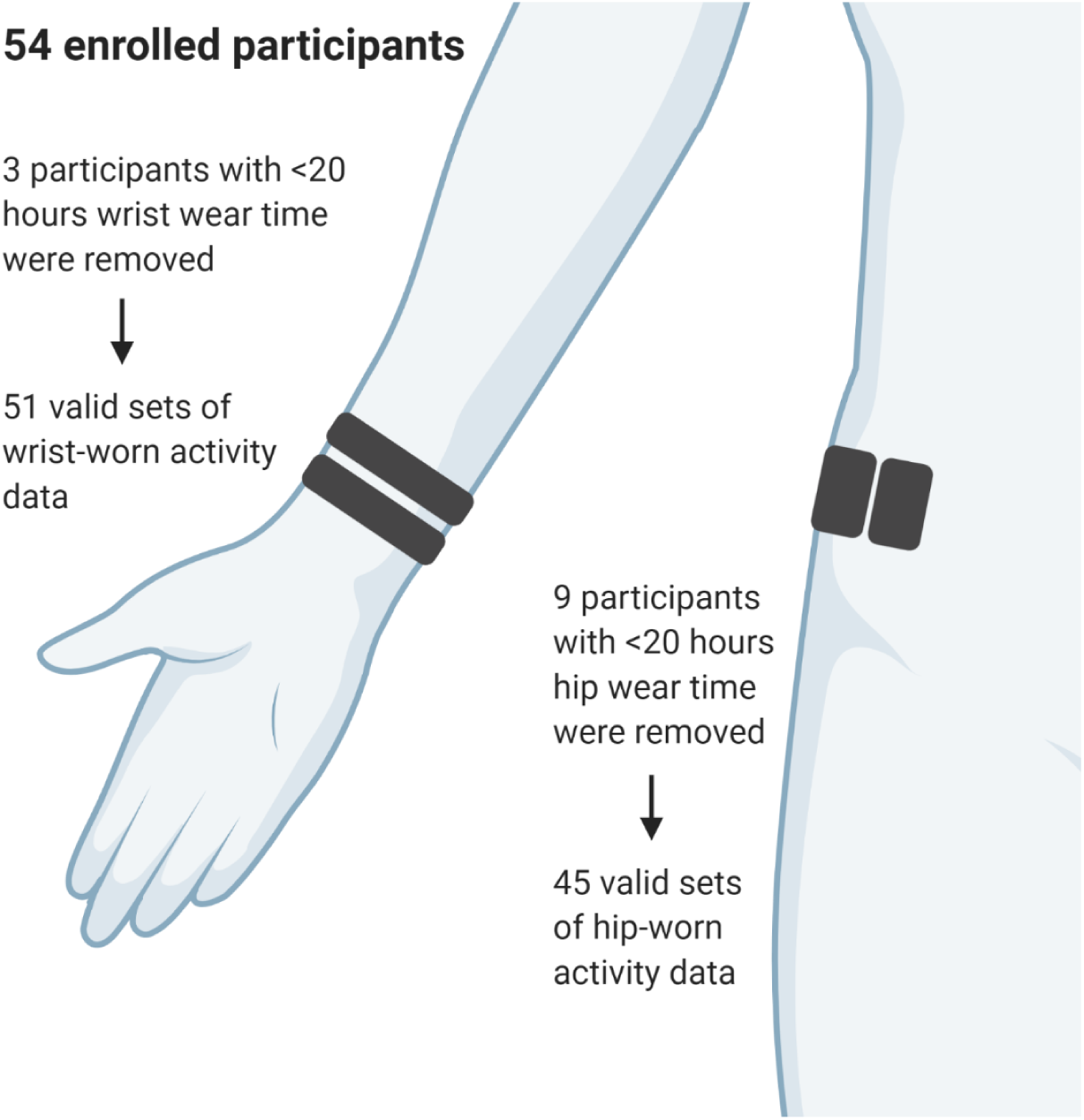
Diagram of accelerometer placement with flow chart of participants with valid wear time at each body location.

Accelerometers were synchronised and programmed via the Open Movement software (v1.0.0.42) (https://openmovement.dev). At each body location, one sensor was programmed to collect data at a sampling rate of 25 Hz while the other collected data at 100 Hz, both with a dynamic range of ±8 *g*. The same four unique sensors were used in all participants. Accelerometer orientation and axis alignment within the hip and wrist straps were consistent with manufacturer guidance and verified prior to the start of each recording session. The assignment of hip or wrist body location and sampling rate of each specific accelerometer was randomized by serial number so that the four accelerometers contributed to measurements at both body locations and sampling rates. Participants additionally filled out an activity diary (Supplement Note 1) to log the times during which they slept, cycled, walked more than 100 meters, ate a meal, participated in self-defined exercise, or removed the accelerometer.

### Acceleration Magnitude

A schematic of our study design is presented in Figure 2. Following the monitoring period, triaxial acceleration data was processed using the open source Biobank Accelerometer Analysis Tool (https://github.com/activityMonitoring/biobankAccelerometerAnalysis) [1]. Acceleration data was automatically calibrated by identifying stationary points within the data to reduce sensor-based measurement bias and offset error [1,15]. Overall wear time was calculated and stationary non-wear episodes of 60 minutes or greater wherein all three axes had a standard deviation of less than 13 m*g* were automatically removed from analysis [1]. Data nominally collected at 25 Hz and 100 Hz were resampled using linear interpolation to generate datasets with precisely 25 and 100 samples per second. A duplicate dataset from the 100 Hz recording was downsampled to 25 Hz for an additional comparison, as implemented in previous studies [13,14]. Vector magnitude of acceleration was calculated as the Euclidean norm of the three accelerometer axes with 1*g* subtracted to account for gravity and negative values truncated to zero. Vector magnitude was collected into 30-second epochs for analysis and overall 24-hour activity was calculated with data reported in milli gravitational units (m*g*) [1].

**Figure 2:**
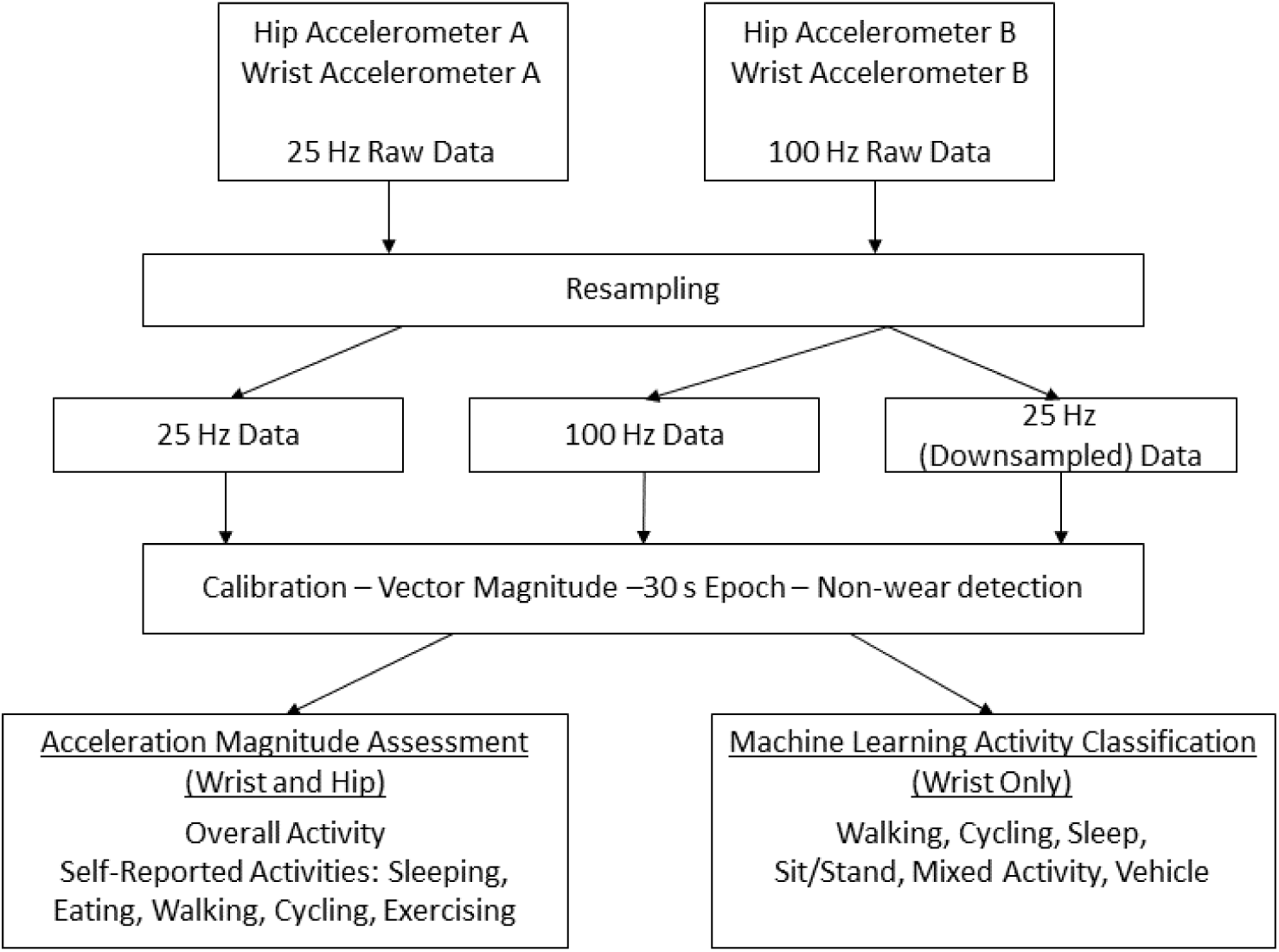
Schematic of study design to inform conversion of accelerometer data collected at 25 Hz to enable comparison with studies collected at 100 Hz.

Vector magnitude was also calculated for all diary-logged bouts of extended walking, cycling, sleeping, exercising, and eating. Time and duration of these activities were extracted from participant activity diaries. Beginning and ending times for diary labels were manually verified by inspecting and cross-referencing raw accelerometer data. If no end time was given for eating activities, a conservative estimate of 15 minutes was designated per meal. Overall moderate-to-vigorous physical activity (MVPA) was additionally quantified, defined as time spent at an acceleration vector magnitude ≥ 100 m*g* at the wrist and ≥ 70 m*g* at the hip [16].

### Activity Classification

Machine learning methods for activity classification of wrist-worn accelerometer data were performed using a two-stage machine learning model of balanced random forests and hidden Markov models [17]. Activities were classified as time spent cycling, mixed activity, sit/stand, sleep, vehicle, and walking. The classification model used in the current study is an open-source model implemented within the Biobank Accelerometer Analysis Tool, having been previously developed and validated for activity classification of the UK Biobank physical activity cohort [17]. The model was trained by Willetts et al., [17] using data from Axivity AX3 accelerometer recording at 100 Hz (±8 g) on the dominant wrist, with concurrent ground truth images recorded every 20 seconds for a total of 160,000 minutes in 134 participants.

### Statistical Analysis

A minimum sample size of 30 participants was calculated to detect a conservative correlation coefficient estimate of 0.6 between sampling rate and body location measurements with 80% power and alpha set at 0.05. This minimum sample size allowed for 30% data loss for dropout, protocol non-compliance, or poor data quality. Participants with less than 20 hours of wear were excluded from analysis. Summary and descriptive statistics were calculated for participant demographics, overall activity levels, and activity levels performed during self-reported activity designations. Two-sided Spearman’s rank correlation was calculated to compare activity levels, time in MVPA, and time in classified activities between accelerometer sampling rates and body locations. Mean difference and relative percentage difference in activity level between sampling rates was calculated with reference to 100Hz measurements. Bland-Altman plots were used to assess fixed or proportional bias and limits of agreement between sampling rates across the full range of participant activity levels. Mean participant activity was plotted in 1-hour increments over a 24-hour day to visualise differences in activity patterns recorded at either sampling rate. Linear regression was used to determine the association between sampling rates and to estimate a conversion factor between acceleration vector magnitude and activity classification recorded at the two sampling rates, with 25 Hz measurements modelled as the predictor, 100 Hz measurements as the regression outcome. Leave-one-subject-out cross validation (*n*-1) was repeated *n* times across each dataset to assess linear model generalisability, with the corresponding root mean square error (RMSE) and R^2^ reported. Statistical analysis was performed in R (v.4.0.0) and RStudio (v1.2.5042).

## Results

### Participant Demographics

Fifty-four healthy adults (33 female, 21 male) with a mean age of 43.4 years (SD 17.6; range 19.5 - 81.2 years) participated in free-living physical activity analysis (Table 1). Acceleration measurement error following self-calibration was less than 2.6 m*g* across all recordings, with no instances of calibration failure or device malfunction. Median hip and wrist accelerometer wear time was 23.7 hours (IQR 0.6) and 24.0 hours (IQR 0.5), respectively. Three sets of wrist-worn and nine sets of hip-worn sensors were removed from analysis as they were worn for less than 20 hours. Participants self-reported an overall mean of 7.4 hours of sleep (SD 1.0), 46.8 minutes of eating (SD 37.8), 28.4 minutes of general exercise (SD 56.7), 58.2 minutes of extended walking (SD 64.0), and 34.1 minutes of cycling (SD 27.8) over the 24-hour assessment period as calculated in participants who logged non-zero time in those activities.

**Table 1:** Participant demographics.

|  | Mean | SD |
| --- | --- | --- |
| Age (years) | 43.4 | 17.6 |
| Height (cm) | 170.5 | 10.8 |
| Weight (kg) | 70.5 | 13.3 |
| BMI (kg/m^2) | 24.2 | 3.6 |
| Sex |  |  |
| Female | 33 | 61% |
| Male | 21 | 39% |
| Handedness |  |  |
| Left | 8 | 15% |
| Right | 46 | 85% |

### 24-Hour Physical Activity Assessment

Mean physical activity across all valid measurements according to time of day is presented in Figure 3 and acceleration data for wrist and hip accelerometers are presented in Table 2. Strong correlation was observed between 25 Hz and 100 Hz recorded data in overall vector magnitude and time spent in MVPA at both the hip (*r* = 0.988) and the wrist (*r* = 0.960 to 0.971). At the hip, vector magnitude collected at 25 Hz resulted in consistently lower overall activity compared to the 100 Hz data, with an absolute mean difference in vector magnitude (95% CI) of 2.0 (1.6, 2.4) m*g* and 9.7 (7.9, 11.5) minutes of MVPA per 24 hours. This represents a −12.4% relative difference in vector acceleration magnitude and −11.2% difference in MVPA time when 25 Hz measurements when compared to 100 Hz measurements at the hip. At the wrist, 25 Hz data collection also resulted in consistently lower overall activity compared to the 100 Hz data, with an absolute mean difference (95% CI) of 4.4 (3.8, 5.0) m*g* in vector magnitude and 25.9 (21.5, 30.3) minutes of MVPA, a −13.3% and −23.1% relative difference in 25 Hz measurements at the wrist, respectively. Bland-Altman plots exhibiting the difference between 25 Hz and 100 Hz 24-hour activity measurements are presented for hip and wrist measurements in Figure 4.

**Table 2:**
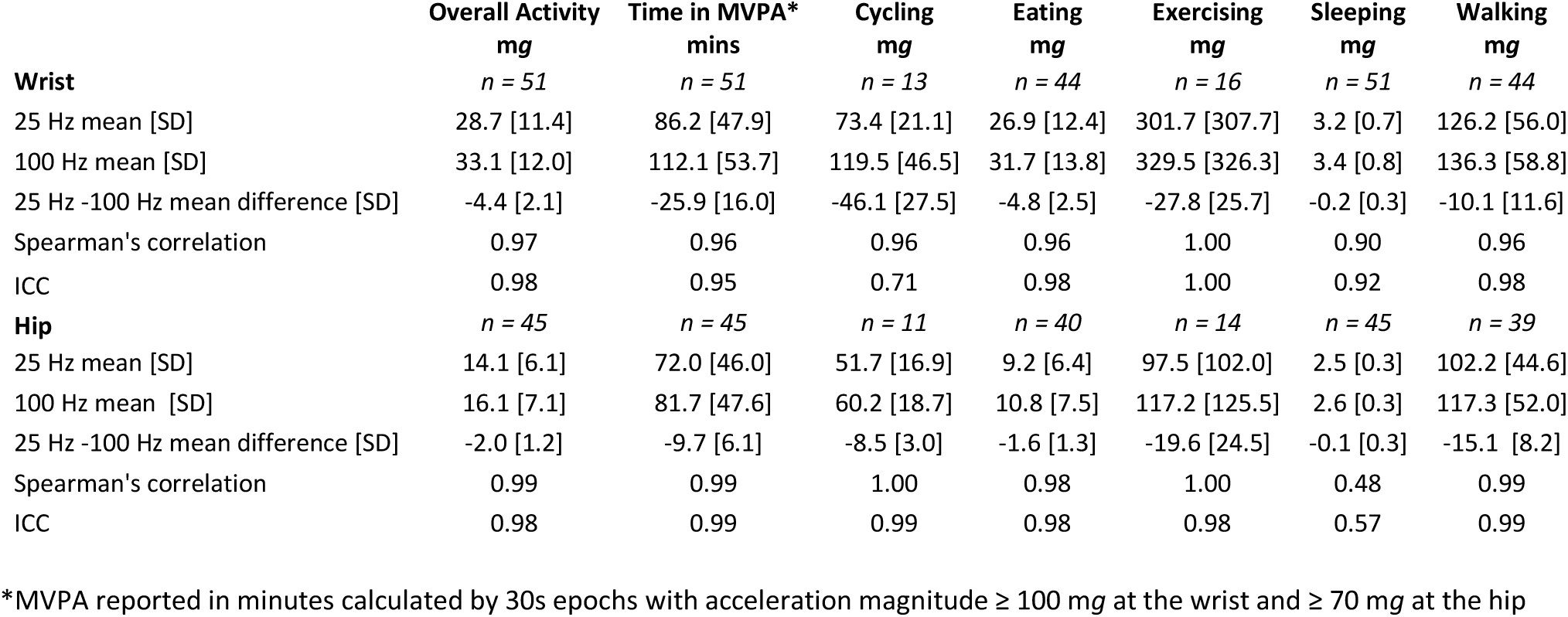
Accelerometer measured physical activity for both overall and diary-reported activities in a convenience sample of 51 adults over a single 24hr period.

|  | Overall Activity<br>mg | Time in MVPA*<br>mins | Cycling<br>mg | Eating<br>mg | Exercising<br>mg | Sleeping<br>mg | Walking<br>mg |
| --- | --- | --- | --- | --- | --- | --- | --- |
| **Wrist** | *n = 51* | *n = 51* | *n = 13* | *n = 44* | *n = 16* | *n = 51* | *n = 44* |
| 25 Hz mean [SD] | 28.7 [11.4] | 86.2 [47.9] | 73.4 [21.1] | 26.9 [12.4] | 301.7 [307.7] | 3.2 [0.7] | 126.2 [56.0] |
| 100 Hz mean [SD] | 33.1 [12.0] | 112.1 [53.7] | 119.5 [46.5] | 31.7 [13.8] | 329.5 [326.3] | 3.4 [0.8] | 136.3 [58.8] |
| 25 Hz -100 Hz mean difference [SD] | -4.4 [2.1] | -25.9 [16.0] | -46.1 [27.5] | -4.8 [2.5] | -27.8 [25.7] | -0.2 [0.3] | -10.1 [11.6] |
| Spearman's correlation | 0.97 | 0.96 | 0.96 | 0.96 | 1.00 | 0.90 | 0.96 |
| ICC | 0.98 | 0.95 | 0.71 | 0.98 | 1.00 | 0.92 | 0.98 |
| **Hip** | *n = 45* | *n = 45* | *n = 11* | *n = 40* | *n = 14* | *n = 45* | *n = 39* |
| 25 Hz mean [SD] | 14.1 [6.1] | 72.0 [46.0] | 51.7 [16.9] | 9.2 [6.4] | 97.5 [102.0] | 2.5 [0.3] | 102.2 [44.6] |
| 100 Hz mean [SD] | 16.1 [7.1] | 81.7 [47.6] | 60.2 [18.7] | 10.8 [7.5] | 117.2 [125.5] | 2.6 [0.3] | 117.3 [52.0] |
| 25 Hz -100 Hz mean difference [SD] | -2.0 [1.2] | -9.7 [6.1] | -8.5 [3.0] | -1.6 [1.3] | -19.6 [24.5] | -0.1 [0.3] | -15.1 [8.2] |
| Spearman's correlation | 0.99 | 0.99 | 1.00 | 0.98 | 1.00 | 0.48 | 0.99 |
| ICC | 0.98 | 0.99 | 0.99 | 0.98 | 0.98 | 0.57 | 0.99 |

\*MVPA reported in minutes calculated by 30s epochs with acceleration magnitude $\geq 100$ mg at the wrist and $\geq 70$ mg at the hip

**Figure 3:**
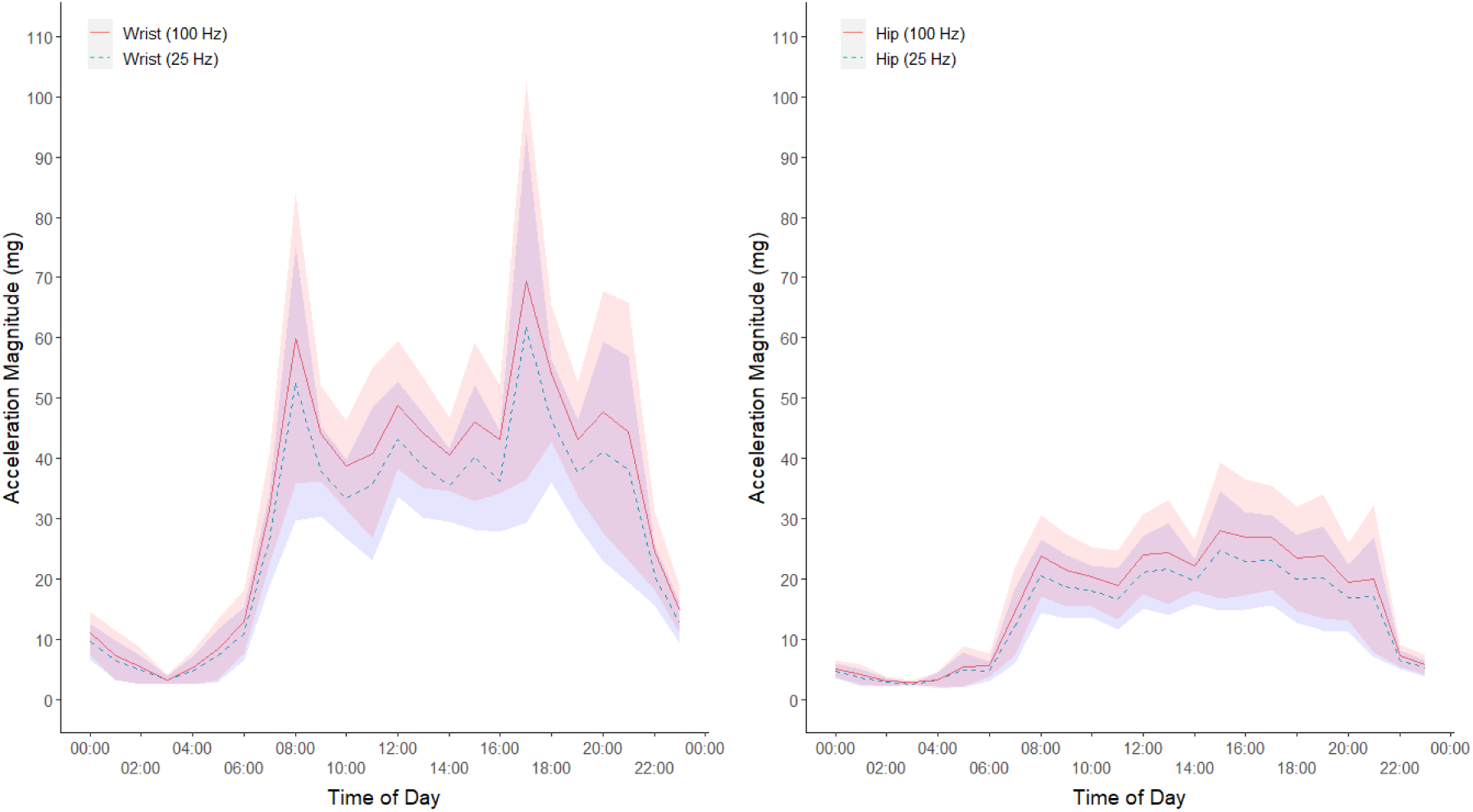
Plot of mean (95%CI) 24-hour activity in 1-hour epochs as measured by acceleration vector magnitude across a convenience sample of 51 participants at each of the four sensor location/rate combinations.

**Figure 4:**
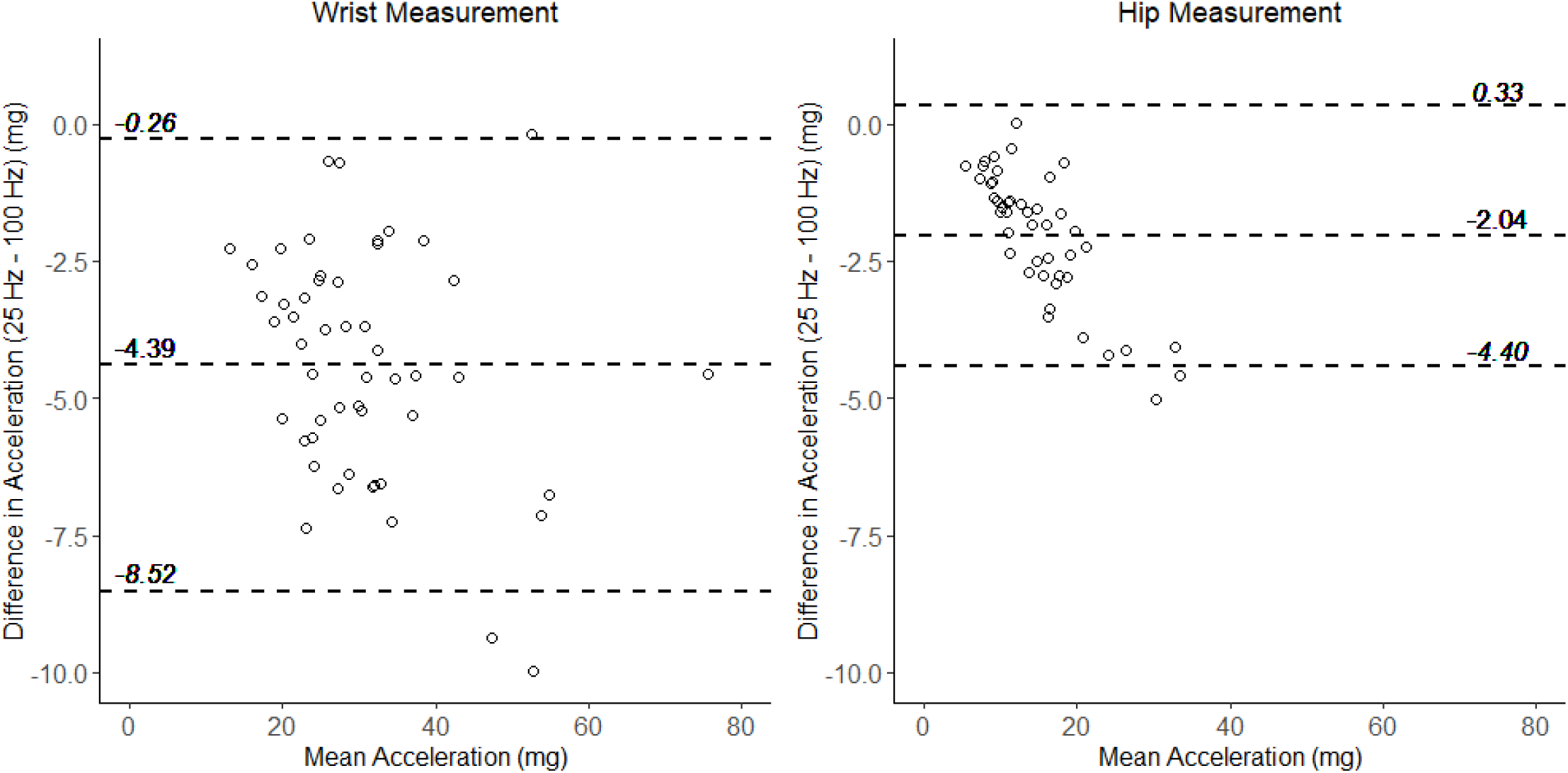
Bland-Altman plots comparing the mean acceleration vector magnitude and differences between the 25 Hz and 100 Hz sensors at the wrist and hip in a convenience sample of 51 adults. Dashed lines indicated mean bias and 95% limits of agreement. Negative bias on the y-axis indicates lower values in the 25 Hz sensor.

At both measurement locations, effectively perfect correlation (r = 1.000) was observed for overall activity between the recorded 100 Hz and the downsampled measurements from the same sensor (Table S1). Comparison of sampling rate via the downsampling of 100 Hz data to 25 Hz in the same sensor resulted in less than 0.1% difference in overall activity and MVPA across both the hip and wrist (Table S1, Figure S1).

### Intensity of Self-Reported Activities

Correlation between acceleration vector magnitude measured at 25 Hz and 100 Hz sampling rates was high during all self-reported non-sleeping activities at the wrist (r = 0.956 to 0.997) and hip (r = 0.977 to 1.000). When recording at the hip, the 25 Hz sampling rate resulted in a mean relative difference in vector magnitude of −12.9% to −16.7% when compared to the 100 Hz sampling rate across all non-sleeping activities. The greatest difference in 25 Hz and 100 Hz data at the wrist was observed during cycling activity, where the 25 Hz accelerometer recorded relative difference of −38.6% when compared to the 100 Hz recording. Other non-sleeping activities resulted in a −7.4% to −15.1% relative difference when recorded at 25 Hz at the wrist. Acceleration during sleep was predictably low, with lower correlation between sampling rates when compared to other activities (r = 0.476 to 0.899). The actual differences are small due to the very activity measured during sleep intervals.

### Machine Learning Activity Classification

Results of the machine learning activity classification are presented in Table 3. Periods of walking, sleep, sit/stand activities, and mixed activities were identified in all participants, whereas periods of cycling and vehicle activities were identified in 13 and 41 participants, respectively. In all participants who had cycling or vehicle activities classified within the 100 Hz 24-hour recording, that activity was also identified within the 25 Hz data. Reduced sampling rate did not produce consistent trends in underreporting of activity across machine learning activity classification as was observed in the quantification of vector magnitude. Activity classification between 25 Hz and 100 Hz measured data was highly correlated (r = 0.855 to 0.967) in all activity categories. The reduced sampling rate resulted in classification of 12.4% (95% CI: 7.5%, 17.4%) more of walking time, 12.8% (95% CI: 6.7%, 18.9%) less time in mixed activities, and 19.2% (95% CI: 8.2%, 30.3%) less vehicle time per participant when compared to 100 Hz collected data in this machine learning model. Cycling, sleep, and sit/stand showed close agreement between sampling rates, with percent differences in time spent in these activities ranging between 0.2% to 4.6%.

**Table 3:** Machine learned activity classification for wrist accelerometers in a convenience sample of 51 adults over a single 24hr period.

|  | Cycling<br>min | Mixed Activity<br>min | Sit/Stand<br>min | Sleep<br>min | Vehicle<br>min | Walking<br>min |
| --- | --- | --- | --- | --- | --- | --- |
| Participants with the identified activity | *n* = 13 | *n* = 51 | *n* = 51 | *n* = 51 | *n* = 41 | *n* = 51 |
| 25 Hz classified mean time [SD] | 27.0 [17.2] | 143.0 [115.0] | 568.4 [139.2] | 475.0 [68.1] | 64.4 [58.6] | 192.4 [91.9] |
| 100 Hz classified mean time [SD] | 28.3 [18.9] | 164.0 [129.6] | 553.3 [148.7] | 475.9 [73.9] | 79.7 [57.2] | 171.1 [89.1] |
| 25-100 Hz mean difference in classified time [SD] | -1.3 [4.0] | -21.0 [36.4] | 15.1 [40.4] | -1.0 [30.8] | -15.3 [28.6] | 21.3 [31.0] |
| Spearman's correlation | 0.97 | 0.92 | 0.95 | 0.86 | 0.89 | 0.92 |
| ICC | 0.98 | 0.96 | 0.96 | 0.91 | 0.88 | 0.94 |

### Converting data collected at lower sample rates to enable comparison with other studies

Results from the linear regression and cross-validation relating sampling rates based on overall 24-hour vector magnitude and activity classification models are presented in Table 4. At the hip and wrist, the linear model of sampling rate accounted for between 97% and 99% of the adjusted variability in the vector magnitude as described by R^2^. Cross-validation across 51 folds at the wrist and 45 folds at the hip resulted in RSME values of 6.5% and 4.7% of the mean wrist and hip vector magnitude, respectively. For each unit of increase in activity of 1 m*g* at the 25 Hz sampling rate, epoch-based vector magnitude at the 100 Hz sample rate increased by 1.158 m*g* (95% CI: 1.120, 1.195) in hip-mounted accelerometers and 1.038 m*g* (95% CI: 0.986, 1.090) in wrist-mounted accelerometers. Acceleration magnitude collected at 25 Hz with a wrist-mounted AX3 accelerometer, Acc_25,_ can be adjusted for comparison to a more standard 100 Hz dataset. Acc_100_, (Box 1) using the relationship: *Acc*_100_ = 1.038(*Acc*_25_) + 3.310. Transformations from 25 Hz to 100 Hz data were generated for mixed, sit/stand, vehicle, and walking activities. In wrist-based activity classification, 69.5% to 92.1% of the variability between sampling rates was accounted for in the models, with cross-validation RMSE representing 7.5% of the mean time in the sit/stand classification and up to 50.5% the mean time spent walking. Due to the small difference in classification of sleep and cycling, no transformation for these activities is recommended.

**Table 4:** Linear regression to convert accelerometer data collected at 25 Hz so that it can be compared to data collected at 100 Hz. R^2^ and RMSE values are reported from Leave-one-subject-out cross-validation.

|  | Coefficient [SE] | 95% CI | p | Intercept [SE] | 95% CI | R<sup>2</sup> | RMSE |
| --- | --- | --- | --- | --- | --- | --- | --- |
| **Acceleration Magnitude (mg)** |  |  |  |  |  |  |  |
| **25 Hz to 100 Hz** |  |  |  |  |  |  |  |
| Wrist | 1.038 [0.026] | (0.986, 1.090) | <0.001 | 3.310 [0.798] | (1.706, 4.914) | 0.967 | 2.155 |
| Hip | 1.158 [0.019] | (1.120, 1.195) | <0.001 | -0.180 [0.283] | (-0.751, 0.391) | 0.988 | 0.757 |
| **Activity Classification at Wrist (min)** |  |  |  |  |  |  |  |
| **25 Hz to 100 Hz** |  |  |  |  |  |  |  |
| Cycling |  |  | Not necessary |  |  |  |  |
| Mixed Activity | 1.085 [0.044] | (0.997, 1.172) | <0.001 | 8.905 [7.952] | (-7.074, 24.88) | 0.921 | 36.1 |
| Sit/Stand | 1.0282 [0.041] | (0.945, 1.111) | <0.001 | -31.14 [24.15] | (-79.67, 17.40) | 0.921 | 41.5 |
| Sleep |  |  | Not necessary |  |  |  |  |
| Vehicle | 0.858 [0.075] | (0.707, 1.009) | <0.001 | 24.47 [6.466] | (11.40, 37.55) | 0.695 | 32.7 |
| Walking | 0.913 [0.047] | (0.820, 1.007) | <0.001 | -4.630 [9.911] | (-24.55, 15.29) | 0.877 | 31.0 |

## Discussion

Outcome measures based on acceleration magnitude, cutpoint thresholds, and activity classification are commonly used in clinical and population studies of physical activity [1,10,18]. To validate the use of reduced sampling rate during physical activity measurement, acceleration magnitude and activity classification was assessed across body location and self-reported activity types. Reduction in recorded sampling rate from 100 Hz to 25 Hz resulted in consistently lower measured overall activity at both hip and wrist body locations using a 30 second epoch mean vector magnitude. Conversely, no consistent pattern in under or overreporting of machine learned activity classification was identified between differing sampling rates. While both acceleration magnitude and activity classification are highly correlated between sampling rates, subtle, yet real differences in outcome measures were observed. Transformation of 25 Hz data can enable extended activity monitoring protocols while retaining comparability to data collected at 100 Hz.

In the current study of healthy adults in the free-living environment, we found a repeatable difference in measured acceleration magnitude and MVPA in a side-by-side comparison of data collection at 25 Hz and 100 Hz. When using a reduced sampling rate a 12% to 13% lower 24-hour activity and 11% to 23% lower time in MVPA was observed, compared to 100 Hz data collection at the hip and wrist. Consistently lower acceleration magnitude was also observed in the 25 Hz recording across diary-logged free-living activities. Similar to the current study, Brønd and Ardivsson [11] found ActiGraph GT3x counts demonstrated small differences between sampling rates (30 Hz, 40 Hz, and 100 Hz) during walking, yet large differences between sampling rates during running, with up to 3000 counts per minute greater intensity recorded in the faster sampling rates. Two other prior investigations of accelerometer sampling rate relied on downsampling accelerometer data from a single sensor, with mixed results and frequently little difference between sampling rates when using raw acceleration-based metrics[13,14]. In this study we conducted a secondary analysis by downsampling data initially collected at 100 Hz, which also resulted in no difference in acceleration magnitude and MVPA at a lower sampling rate, even while raw data collection at 25 Hz demonstrated a reduction in the activity intensity captured during acceleration analysis. Subsequently, basic downsampling of data from a single accelerometer is probably insufficient to determine the full effect of different sampling rates.

When using machine learning models of activity classification, we found high correlation between different sampling rates across all classified activities. No repeatable pattern in under or over reporting was observed in the reduced sampling rate, however, higher walking time and lower vehicle travel time classified in the 25 Hz data. This finding is contrary to a study published by Zhang et al. [12] which reported no significant reduction in activity classification accuracy when comparing downsampled data to the original 80 Hz sequence. It is likely the use of downsampling versus a side-by-side comparison has contributed to the opposing findings. Compared to overall vector magnitude and the sit/stand classification, higher RMSE relative to mean values were observed in transformation model for classified time in mixed activity, vehicle, and walking. This may be the result of fewer participants and less time spent in these activities within the current dataset relative to low vector-magnitude sit/stand activities.

Most recent physical activity studies include protocols where acceleration data is collected at a sampling frequency between 20 and 100 Hz [10,18,19]. It is important to note that reporting of sampling rate used in testing protocols remains a problem in the field of physical activity, with recent reviews finding that 16% to 73% studies failed to report the sampling rate used in their data collection protocol [10,18]. In an assessment of benchmark datasets, Khan et al [20] have argued that typical accelerometer sampling rates in many human motion studies are up to 57% higher than necessary for adequate data analysis, with optimal sampling rates between 12 and 63Hz depending on body location and model of accelerometer. Migueles et al[18], however, argue that future data processing needs are unknown, and thus the highest possible sampling frequency should be used. The world’s largest objectively-measured physical activity database contains accelerometer data from 100 Hz data collection using the AX3 device [1]. Based on the results of the current study, if a reduced sampling rate of 25 Hz is selected in a measurement protocol, acceleration magnitude can be adjusted for comparison to a more standard 100 Hz dataset.

### Strengths and limitations

A major strength of our study is that it includes a side-by-side comparison of standard and reduced accelerometer sampling rates at multiple body locations, without dependency on downsampling. This methodology includes the comprehensive use of vector magnitude, cut-point, and machine learning variables to facilitate direct comparison of data collected at 25 Hz with existing large physical activity cohorts that have collected data at 100 Hz. However, data was collected in a healthy convenience sample, and there may be unquantified effects of measurement in disease populations. While individual devices were randomised to body location (hip/wrist), a potential limitation is that the exact position of devices at each location was not randomised by design. For example, when sensors were placed side-by-side on participants’ wrists, it could be possible that the 100 Hz sensor was more frequently placed nearer to the wrist and thus experience slightly higher accelerations. However, we feel this alternative explanation is unlikely to account for the differences we and others have found [11]. Epoch-based comparisons can have sensitivity to exact epoch boundaries, as all individual sensor clocks drift slightly over time. Comparing two epochs between devices that report the same boundaries may be comparing two slightly different periods.

## Conclusion

Reducing accelerometer sampling rate from 100 Hz to 25 Hz can dramatically extend study monitoring periods while still providing valid data, after appropriate transformations. In order to facilitate comparisons between studies, researchers should consider and discuss the effect of sampling rate on data analysis, and fully report all accelerometer parameters in the study methodology.

## Supporting information

STROBE

ICMJE

ICMJE

ICMJE

ICMJE

ICMJE

ICMJE

ICMJE

## Data Availability

Following an embargo period, data will be available from the corresponding author upon reasonable request.

## ACKNOWLEDGEMENTS

SS is supported by the Clarendon Fund from Oxford University and the Oxford University Press. SK, AD and PD are supported by the National Institute for Health Research (NIHR) Oxford Biomedical Research Centre (BRC). DJ is a director and shareholder of Axivity, Ltd. AD is supported by Health Data Research UK. AD and SC are supported by the Alan Turing Institute and the British Heart Foundation (grant number SP/18/4/33803). The views expressed are those of the author(s) and not necessarily those of the NIHR. Funding sources played no role in study design. All authors have completed ICMJE disclosure forms, which are included in the supplemental material. The STROBE checklist for cohort studies is also included in the supplemental material. Figure 1 was created with BioRender.com.

## Box 1

**Example of converting data collected at 25 Hz so that it can be compared to data collected at 100 Hz**

A 68 year-old female participant has 7-day mean overall acceleration magnitude of 24.8 m*g* as recorded at 25 Hz on the dominant wrist with an AX3 accelerometer. Researchers wish to compare this individual to the baseline 65-74 year-old female in the UK Biobank database 26.6 (SD 7.1) m*g*, recorded at 100 Hz with an AX3 accelerometer[1]. Without sampling rate conversion, a 24.8 m*g* activity level is below average for her age group, and places this patient in the 40th percentile of activity in the 65-74 year-old female cohort. With 25 Hz to 100 Hz conversion following the wrist-based regression model, Acc_100_ = 1.038*Acc_25_ + 3.310, the participant’s comparable acceleration magnitude is 29.1 m*g*, placing her in the 64th percentile of the appropriate demographic cohort.

