## Supplementary material for "Impact of Reduced Sampling Rate on Accelerometer-based Physical Activity Monitoring and Machine Learning Activity Classification": Figure S1

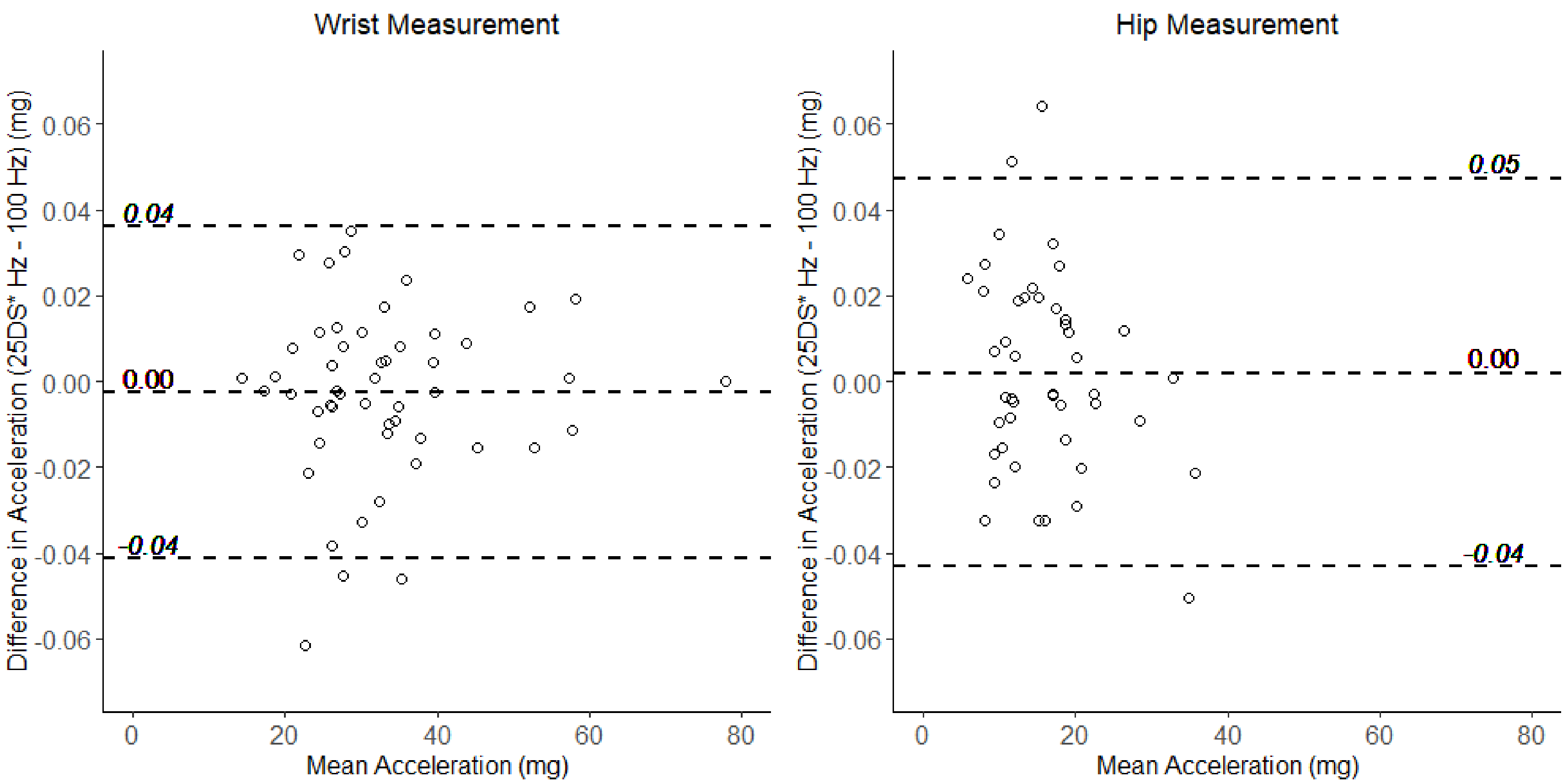


**Supplemental Figure 1:** Bland-Altman plots comparing the mean acceleration vector magnitude and differences between the downsampled 25 Hz acceleration and 100 Hz raw acceleration from sensors recording at 100 Hz at the wrist and hip. Dashed lines indicated mean bias and 95% limits of agreement. Negative bias on the y-axis indicates lower values in the downsampled 25 Hz data.
