## Supplement Note 1 for "Impact of Reduced Sampling Rate on Accelerometer-based Physical Activity Monitoring and Machine Learning Activity Classification"

### Activity and Sleep Diary

Instructions: Please complete the diary to record physical activity that you complete, while wearing your activity monitors, during your 24-hour activity monitoring session. Please note the beginning and ending time of each activity to the nearest 5 minutes, if possible.

| | STUDY DAY 1 | STUDY DAY 2 |
|---|---|---|
| What time did you go to bed? | | |
| What time did you wake up? | | |
| At what times, if any, did you ride a bicycle? | | |
| At what times, if any, did you take a walk (more than 100 meters)? | | |
| At what times did you do any other exercise (please note specific activity)? | | |
| What times did you eat meals? | | |
| At what times, if any, did you remove the monitors? | | |
