## Supplementary material for "Impact of Reduced Sampling Rate on Accelerometer-based Physical Activity Monitoring and Machine Learning Activity Classification": Table S1

**Supplemental Table 1: Overall and Diary-Associated Acceleration Vector Magnitude and MVPA - Comparison Between 100 Hz and 25 Hz-Downsampled Data**

| | **Overall Activity m*g*** | **Time in MVPA** mins** | **Cycling m*g*** | **Eating m*g*** | **Exercising m*g*** | **Sleeping m*g*** | **Walking m*g*** |
|---|---|---|---|---|---|---|---|
| **Wrist** | *n = 51* | *n = 51* | *n = 13* | *n = 44* | *n = 16* | *n = 51* | *n = 44* |
| 100 Hz mean [SD] | 33.1 [12.0] | 112.1 [53.7] | 119.5 [46.5] | 31.7 [13.8] | 329.5 [326.3] | 3.4 [0.8] | 136.3 [58.8] |
| 25 Hz DS* [SD] | 33.1 [12.0] | 112.1 [53.6] | 119.3 [45.9] | 31.7 [13.8] | 329.5 [326.2] | 3.4 [0.9] | 136.3 [58.7] |
| 25 DS*-100 mean absolute difference [SD] | 0.0 [0.1] | -0.0 [1.0] | -0.2 [1.1] | 0.0 [0.1] | 0.0 [0.2] | 0.0 [0.0] | 0.0 [0.1] |
| Spearman's correlation coef | 1.00 | 1.00 | 1.00 | 1.00 | 1.00 | 1.00 | 1.00 |
| ICC | 1.00 | 1.00 | 1.00 | 1.00 | 1.00 | 1.00 | 1.00 |
| **Hip** | *n = 45* | *n = 45* | *n = 11* | *n = 40* | *n = 14* | *n = 45* | *n = 39* |
| 100 Hz mean [SD] | 16.1 [7.1] | 81.7 [47.6] | 60.2 [18.7] | 10.8 [7.5] | 117.2 [125.5] | 2.6 [0.3] | 117.3 [52.0] |
| 25 DS*Hz [SD] | 16.1 [7.1] | 81.8 [47.6] | 60.2 [18.7] | 10.8 [7.5] | 117.2 [125.7] | 2.6 [0.3] | 117.3 [52.0] |
| 25 DS*-100 mean absolute difference [SD] | 0.0 [0.0] | 0.0 [0.6] | 0.0 [0.1] | 0.0 [0.0] | 0.1 [0.2] | 0.0 [0.0] | 0.0 [0.2] |
| Spearman's correlation coef | 1.00 | 1.00 | 1.00 | 1.00 | 1.00 | 1.00 | 1.00 |
| ICC | 1.00 | 1.00 | 1.00 | 1.00 | 1.00 | 1.00 | 1.00 |

*DS – Recorded at 100 Hz and downsampled to 25 Hz

**MVPA reported in minutes calculated by 30s epochs with acceleration magnitude >100 m*g* at the wrist and >70 m*g* at the hip
